# Seroprevalence of Coronavirus Disease 2019 (COVID-19) Among Health Care Workers from Three Pandemic Hospitals of Turkey

**DOI:** 10.1101/2020.08.19.20178095

**Authors:** Gizem Alkurt, Ahmet Murt, Zeki Aydin, Ozge Tatli, Nihat Bugra Agaoglu, Arzu Irvem, Mehtap Aydin, Ridvan Karaali, Mustafa Gunes, Batuhan Yesilyurt, Hasan Turkez, Adil Mardinoglu, Mehmet Doganay, Filiz Basinoglu, Nurhan Seyahi, Gizem Dinler Doganay, Levent Doganay

**Author notes:** **Corresponding authors**: Levent DOGANAY, MD. Saglik Bilimleri Universitesi, Umraniye Egitim ve Arastirma Hastanesi, Gastroenteroloji ve Hepatoloji Klinigi, Istanbul, Turkey., Gizem DINLER DOGANAY, PhD. Istanbul Teknik Universitesi, Molekuler Biyoloji ve Genetik Bolumu, Istanbul, Turkey., Nurhan SEYAHI, MD. Istanbul Universitesi, Cerrahpasa Tip Fakultesi, Dahili Tip Bilimleri Bolumu, Nefroloji Bilim Dali, Istanbul, Turkey., Filiz BASINOGLU, MD. Darica Farabi Egitim ve Arastirma Hastanesi, Tibbi Biyokimya Bolumu, Kocaeli, Istanbul, Turkey. These authors contributed to the study equally (Gizem Alkurt, Ahmet Murt, Zeki Aydin, Ozge Tatli). These authors have equal correspondence (Filiz Basinoglu, Nurhan Seyahi, Gizem Dinler Doganay, Levent Doganay).

## Abstract

COVID-19 is a global threat with an increasing number of infections. Research on IgG seroprevalence among health care workers (HCWs) is needed to re-evaluate health policies. This study was performed in three pandemic hospitals in Istanbul and Kocaeli. Different clusters of HCWs were screened for SARS-CoV-2 infection. Seropositivity rate among participants was evaluated by chemiluminescent microparticle immunoassay. We recruited 813 non-infected and 119 PCR-confirmed infected HCWs. Of the previously undiagnosed HCWs, 22 (2.7%) were seropositive. Seropositivity rates were highest for cleaning staff (6%), physicians (4%), nurses (2.2%) and radiology technicians (1%). Non-pandemic clinic (6.4%) and ICU (4.3%) had the highest prevalence. HCWs in “high risk group” had similar seropositivity rate with “no risk” group (2.9 vs 3.6 *p*=0.7), indicating the efficient implementation of protection measures in the hospitals in Turkey. These findings might lead to the re-evaluation of infection control and transmission dynamics in hospitals.

## INTRODUCTION

In late 2019, a novel coronavirus (SARS-CoV-2) has emerged in Wuhan, China and posed a global threat to public health with a quick spread and escalating mortality. As of June, 2020, SARS-CoV-2 related disease COVID-19 affected more than nine million people worldwide, and caused more than 500 thousand deaths (WHO, 2020). This is the third coronavirus outbreak that the world has faced in the last two decades, and it apparently will not be the last.

Typical initial clinical signs of COVID-19 have been reported as fever, dry cough, fatigue, headache and shortness of breath (Guan et al., 2020). Less commonly, diarrhea, nausea and vomiting were also reported as the atypical symptoms of the disease (Wang et al., 2020b). Older age and comorbid conditions, particularly hypertension and diabetes increase hospitalization and mortality rates among infected individuals. Unknown percentage of asymptomatic carriers is another concern due to the difficulty of tracing their contacts with possible inaccurate calculations of R-naught (Park et al., 2020; Yu and Yang, 2020).

Based on genome sequence analysis, SARS-CoV-2 genome was reported to contain 14 Open Reading Frames (ORFs) encoding 27 proteins (Wu et al., 2020). The lipid envelope of the virus possesses primarily three structural proteins including membrane (M), envelope (E), and spike (S) proteins. Both S and N proteins have been reported as highly abundant and immunogenic, which makes them potential targets for serological diagnosis (Okba et al., 2020). Besides the viral nucleic acid detection based on real-time reverse transcription polymerase chain reaction (real-time RT-PCR)(Shih et al., 2020), immunoassay tests were recently developed for accurate detection of IgG and IgM antibodies against SARS-CoV-2 in sera samples. Such tests may help to eliminate false negatives of RT-PCR tests caused by the difference in the viral load of different respiratory specimens (Liu et al., 2020). For increased sensitivity in diagnosis of COVID-19, both modalities should be combined as complementary to each other. Moreover, serologic tests could be utilized for the detection of overall infection rate in the population (Lou et al., 2020).

Istanbul is the epicenter of the ongoing pandemic in Turkey and 60% of the confirmed cases are from Istanbul (Doganay et al., 2020). In the current study, seroprevalence of COVID-19 specific IgGs was tested among health care workers (HCWs) from two different pandemic hospitals in Istanbul and one from the neighboring city of Kocaeli. First COVID-19 case was officially reported on 11 March 2020 in Turkey and since then all three hospitals had a substantial role with treating more than 40 000 patients during pandemic. We aimed to identify asymptomatic infections and to assess the risks of contracting COVID-19 among different clusters of HCWs. Evaluating the prevalence of infection at different parts of the hospitals and the effect of transmission prevention measures hold great importance for the development of mitigation strategies.

## RESULTS

### Demographic data

Locations of the three hospitals in Istanbul and Kocaeli were mapped in Figure 1. The timeline showing the progression of COVID-19 pandemic in Turkey, which includes events from the date that the country’s first coronavirus case has been announced, was given in Figure 2. All three pandemic hospitals are tertiary health care centers with high capacity, employing a total of 8328 HCWs. On the peak day of the pandemic, maximum hospitalized patient numbers (on a day) reached to 410, 222 and 300 for UEAH, Cerrahpasa and Farabi, respectively. Total number of hospitalized patients in the hospitals throughout the pandemic was 5437 (Table 1). Among 8328 HCWs, 932 were enrolled for the study. Demographics, assigned work areas during pandemic, comorbidities and SARS-CoV-2 IgG positivity according to risk stratification of HCWs were given on Table 2. Of all participants, the number of personnel classified as “no risk”, “low risk” and “high risk” group were 113 (12.1%), 157 (16.8%) and 543 (58.3%), respectively. Additional 119 HCWs (12.8% of all participants), previously diagnosed with COVID-19 by RT-PCR, were enrolled and classified as “PCR positive” group. Of PCR positive HCWs, 103 displayed the clinical signs of COVID-19. 16 (13.4%), who did not have compatible symptoms, were tested due to the exposure history, and were recorded as asymptomatic. Only one HCW had a history of admission to ICU, no deaths occurred (Table 1). Of the non-infected HCWs, 597 gave consent for oro-nasopharyngeal swab sampling and all RT-PCR results were negative.

**Figure 1:**
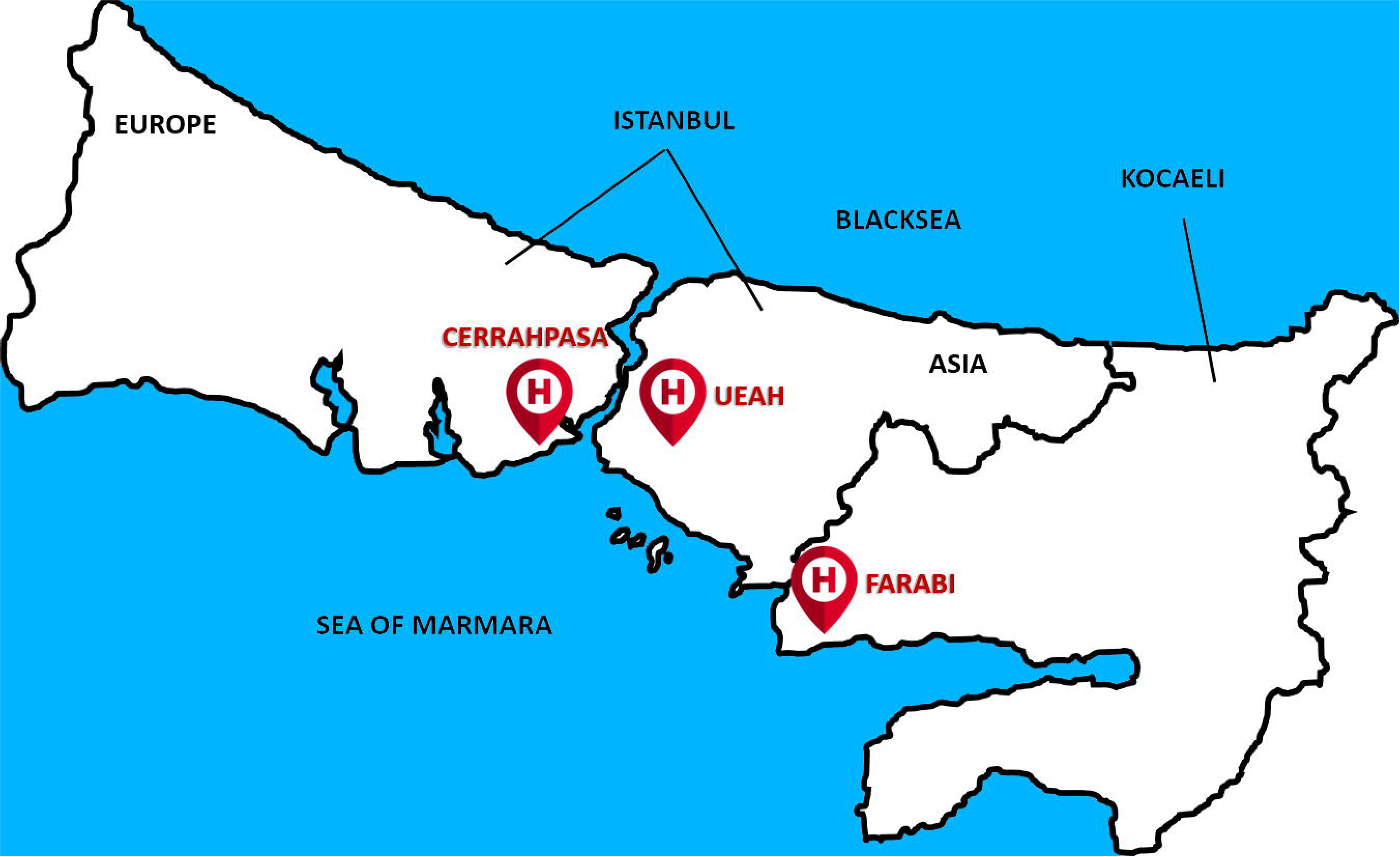
Location of three pandemic hospitals

**Figure 2:**
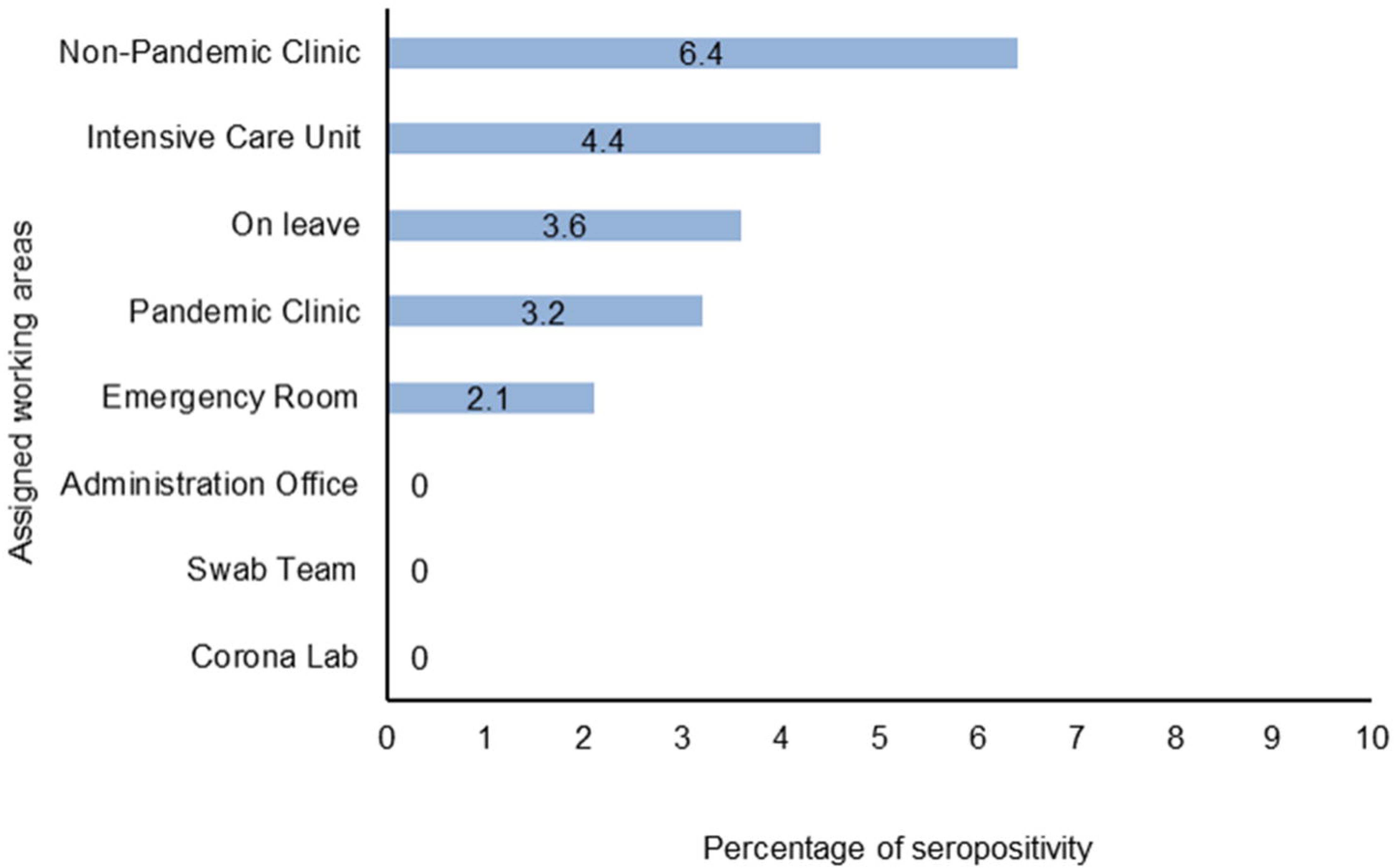
Timeline of COVID19 in three pandemic hospital

**Table 1.** Characteristics of the Health Care Facilities

|  | Hospitals |  |  | Total |
| --- | --- | --- | --- | --- |
|  | UEAH | Cerrahpasa | Farabi |  |
| Number of Hospital Beds | 836 | 880 | 350 | 2066 |
| Maximum Number of Hospitalized Patient with COVID-19 (at the peak day) | 410 | 222 | 300 | 932 |
| Total Number of Hospitalized Patient with COVID-19 | 2528 | 1056 | 1853 | 5437 |
| Total Number of Patients Visits at COVID-19 Outpatient Clinic | 14906 | 21970 | 12375 | 49251 |
| Number of HCWs | 3232 | 3723 | 1373 | 8328 |
| HCWs diagnosed with COVID-19 | 125<br>(3.8%) | 107<br>(2.8%) | 14<br>(1%) | 241<br>(2.8%) |
| PCR Positive HCWs | 105<br>(3.2%) | 84<br>(2.2%) | 8<br>(0.5%) | 187<br>(2.2%) |
| Hospitalized HCWs in ICU | 1 | 0 | 0 | 1 |
| Number of Deaths in infected HCWs | 0 | 0 | 0 | 0 |

All data is given for the period between 1 March and 30 May 2020. HCW: Health Care Worker

**Table 2:**
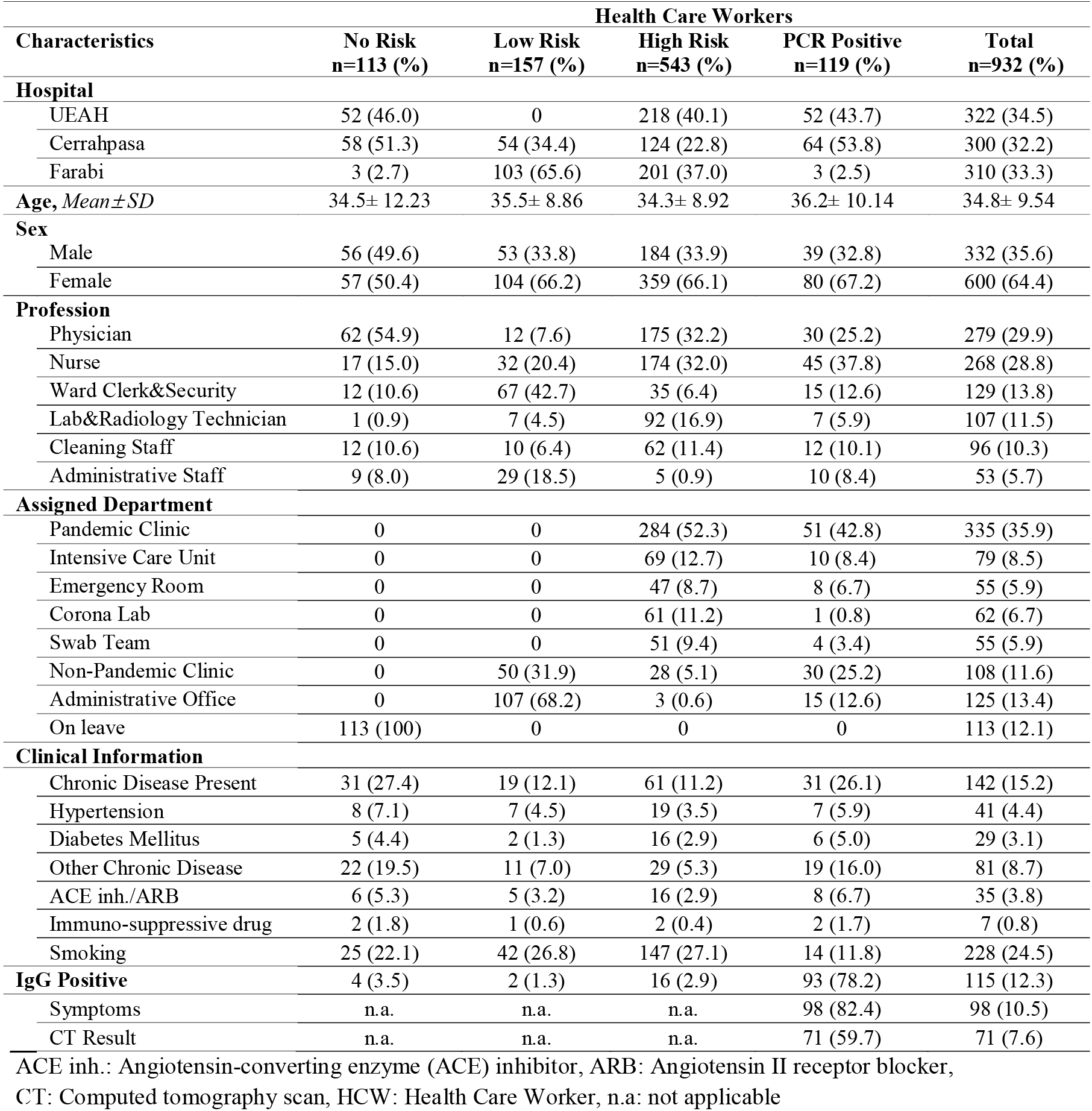
Demographics and seropositivity of front-line and non-front-line HCWs

### Seroprevalence of HCWs in non-infected group

IgG antibodies against SARS-CoV-2 in serum samples of all participants were detected by chemiluminescent microparticle immunoassay. The rate of seroprevalence was 2.7% among non-infected HCWs (Table S2). The seropositivity rate was 3.5%, 1.3%, 2.9% in “no risk”, “low risk” and “high risk” groups, respectively (*p*=0.4). Among three hospitals, Cerrahpasa had the highest seroprevalence (7.2%, *p*<0.001) (Table 3/Figure S1). The seropositivity rate in Cerrahpasa was statistically significantly different when compared to UEAH (*p*<0.01) and Farabi (*p*<0.001). Of 307 HCWs in Farabi, only one tested positive, yielding the lowest seropositivity rate of the hospitals (0.3%) (Figure 3). Although the seroprevalence was not significantly different among professions, it was highest among cleaning staff (6%) (Figure 4). Interestingly, the seropositivity rate of the workers employed in non-pandemic clinics (6.4%) was higher than those working in other areas (*p*=0.05) (Table 4/Figure 5).

**Table 3:** Seropositivity among risk groups

| Characteristic | No Risk<br>n=113 (%) | Low Risk<br>n=157 (%) | High Risk<br>n=543 (%) | Total<br>n=813 (%) |
| --- | --- | --- | --- | --- |
| **IgG Positive** | 4 (3.5) | 2 (1.3) | 16 (2.9) | 22 (2.7) |
| **Hospital** |  |  |  |  |
| UEAH | 1 (1.9) | N/A | 3 (1.3) | 4 (1.5) |
| Cerrahpasa | 3 (5.1) | 2 (3.7) | 12 (9.6) | 17 (7.2) |
| Farabi | 0 | 0 | 1 (0.4) | 1 (0.3) |
| **IgG Titration, mean±SD** | 0.4 ±0.18 | 0.3 ±0.11 | 0.1 ±0.16 | 0.3 ±0.16 |
| N/A: Not Available |  |  |  |  |

**Figure 3:**
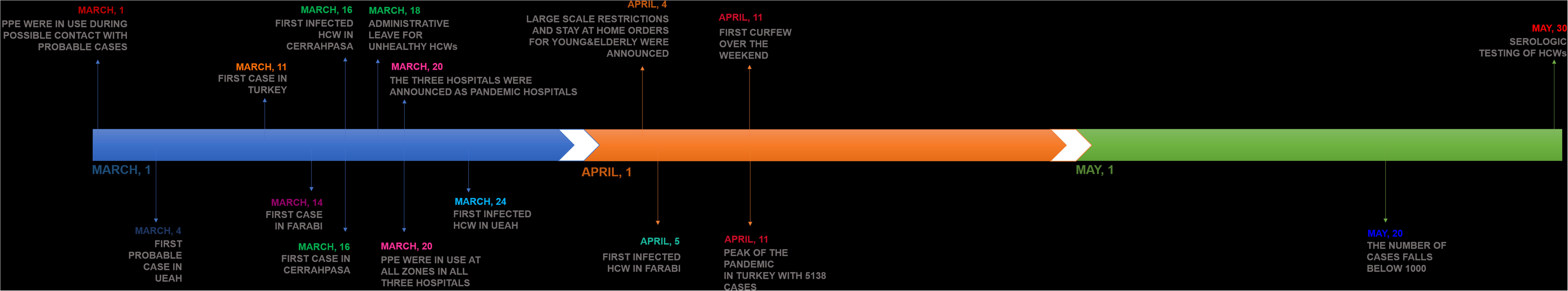
Seropositivity results of non-infected HCW in three pandemic hospitals

**Figure 4:**
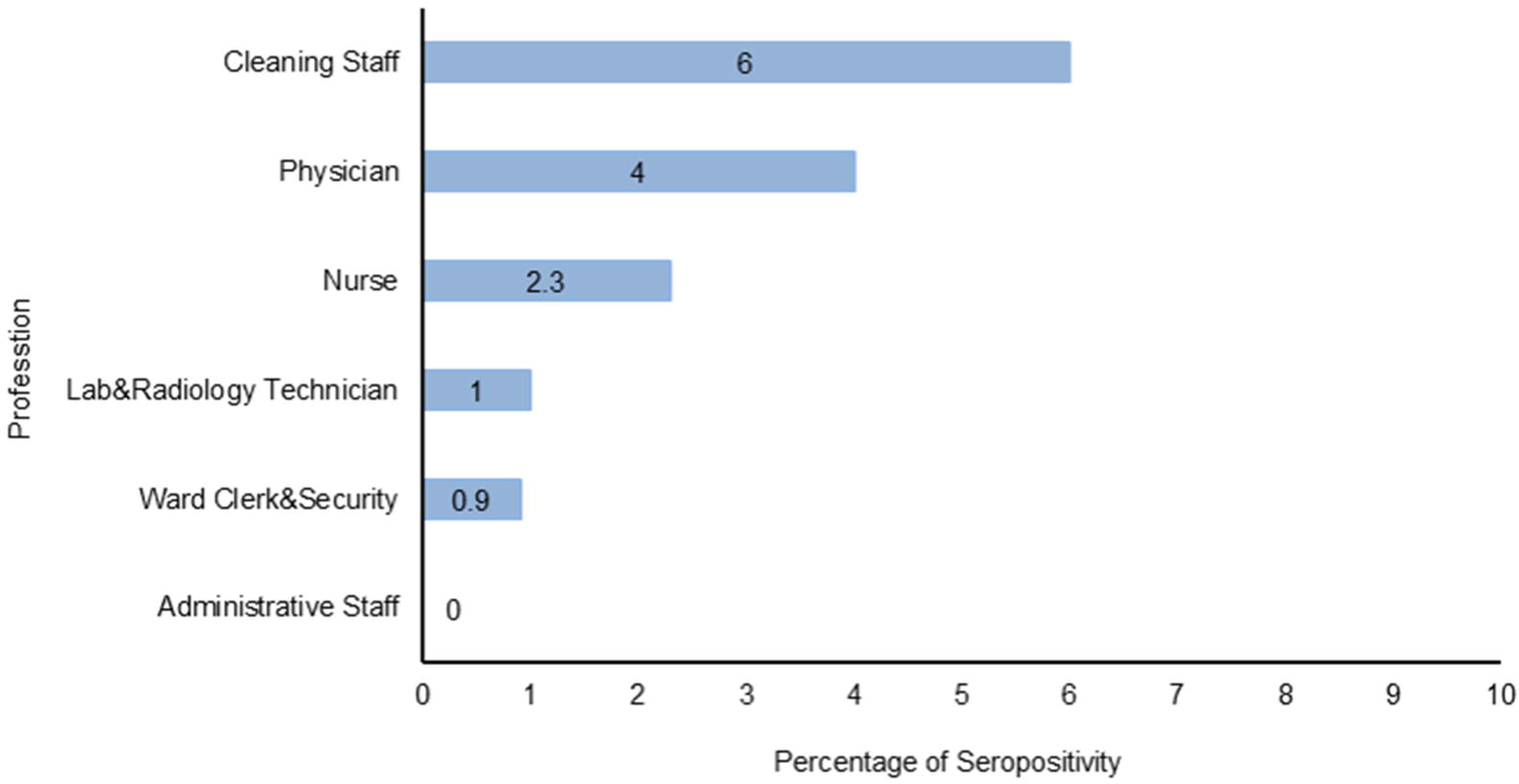
Seropositivity results of non-infected HCW according to profession

**Table 4:** Seropositivity among assigned working areas and profession

| Characteristic<br>n (%) | IgG Negative<br>n=791 | IgG Positive<br>n=22 | OR | 95% CI | p value |
| --- | --- | --- | --- | --- | --- |
| **Profession** |  |  |  |  |  |
| Physician | 239 (96.0) | 10 (4.0) | 1.93 | 0.82- 4.51 | 0.16 |
| Nurse | 218 (97.7) | 5 (2.2) | 0.77 | 0.28-2.12 | 0.81 |
| Ward Clerk&Security | 113 (99.1) | 1 (0.9) | 0.29 | 0.04-2.15 | 0.35 |
| Lab&Radiology Technician | 99 (99.0) | 1 (1.0) | 0.33 | 0.04-2.50 | 0.51 |
| Cleaning Staff | 79 (94.0) | 5 (6.0) | 2.65 | 0.95-7.38 | 0.07 |
| Administrative Staff | 43 (100) | 0 | n.a. | n.a. | 0.62 |
| **Assigned Department** |  |  |  |  |  |
| Pandemic Clinic | 276 (96.8) | 9 (3.2) | 1.3 | 0.55-3.08 | 0.65 |
| Intensive Care Unit | 66 (95.6) | 3 (4.3) | 1.73 | 0.5-6.01 | 0.42 |
| Emergency Room | 46 (97.9) | 1 (2.1) | 0.77 | 0.10-5.86 | 1.00 |
| Corona Lab | 61 (100) | 0 | n.a. | n.a. | 0.4 |
| Swab Team | 51 (100) | 0 | n.a. | n.a. | 0.63 |
| Non-Pandemic Clinic | 73 (93.6) | 5 (6.4) | 2.89 | 1.04-8.61 | 0.05 |
| Administration Office | 111 (100) | 0 | n.a. | n.a. | 0.06 |
| On leave | 107 (96.4) | 4 (3.6) | 1.39 | 0.46-4.19 | 0.53 |

n.a: not applicable, OR: Odd Ratio, CI: Confidence Interval

**Figure 5:**
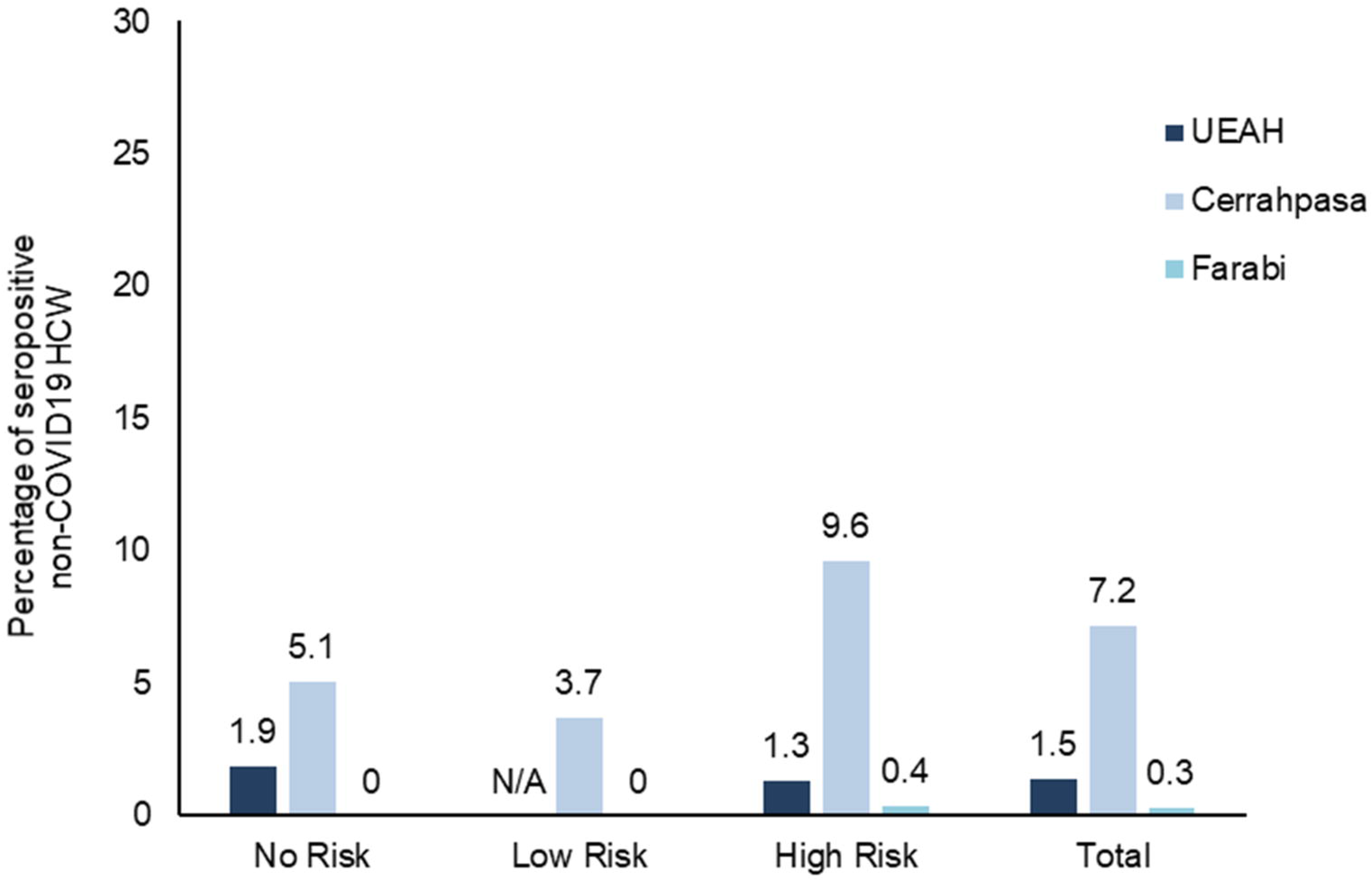
Seropositivity results of non-infected HCW according to assigned working areas

### Seroprevalence of PCR positive group

To assess antibody production in COVID-19 patients, we analyzed the positive rates of IgGs in sera of all HCWs after 52.8±11.6 days post-infection. IgGs for SARS-CoV-2 were detected in 78.2% of convalescent COVID-19 patients (Table S1). Among PCR positive group, those had CT findings compatible with COVID-19, had higher seropositivity (*p*<0.001). IgG titers in asymptomatic PCR positive patients were significantly lower than symptomatic ones (*p*=0.008). There were 10 HCWs with IgG titers between 1.4–1.0 S/C. If the cut off was set to 1.0 S/C, seropositivity in this risk group would have increased to 82.3%.

## DISCUSSION

Health care systems are under tremendous pressure due to the lack of curative treatment for COVID-19.((Wang et al., 2020a) Protection of first-line HCWs from the infection is of utmost importance to provide sustainable public health services. High burden of COVID-19 disease in hospitals worldwide has been explored in several studies. In Spain, one of the European countries hardest-hit by COVID-19, the nationwide seroprevalence for SARS-CoV-2 tested by chemiluminescent microparticle immunoassay was found to be around 5%. For HCWs, the seroprevalence has surpassed the national rate and reached over 8%.(Pollán et al., 2020) Of HCWs in a tertiary hospital in Belgium, seroprevalence detected by rapid cassette test was 6.4%.(Steensels et al., 2020) In another study conducted in Wuhan, 1.1% of HCWs tested positive in their swab samples by RT-PCR (Lai et al., 2020). 0.89% of HCWs in two hospitals in Netherlands had positive RT-PCR results for COVID-19 (Kluytmans-van den Bergh et al., 2020). In this study, even though the percentage of HCWs infected with COVID-19 in the three pandemic hospitals is also noteworthy (2.7%), the insignificant difference between no-risk and high-risk group implied that the protection measures are reassuringly rigorous to prevent the transmission of SARS-CoV-2 in these hospitals.

HCWs employed in coronavirus testing labs (Corona lab) and swab teams were reasonably anticipated to be at high risk. In this study, we did not observe seropositivity in these personnel, they were found to be efficiently protected from the disease. However, non-clinical healthcare workers such as cleaning staff were found to be more prominently affected from the transmission possibly due to the inadequate compliance with the infection control. Thus, all staff in hospitals should be well-trained on elements of disease transmission; such as the sources of exposure to the virus, risks associated with the exposure and suitable occupational protocols. Besides, the workers in non-pandemic clinics were apparently exposed to the virus regardless of specific exposure risk. Such data implied that the risk of viral transmission in these areas are widely underestimated and utmost caution is urged in all zones of the hospitals.

Limited data are available for asymptomatic or subclinical infections in transmission of SARS-CoV-2 virus (Weitz et al., 2020). Here, 13.4% of the PCR-confirmed HCWs with infection never developed any COVID-19 relevant symptoms and remained asymptomatic. Besides, a substantial number of undiagnosed HCWs were seropositive indicating the recovery from COVID-19 without any clinical signs of the disease. These asymptomatic carriers who remained undiagnosed throughout the infection may be the silent sources of virus spread among HCWs. The finding that IgG levels of asymptomatic individuals were lower than that of symptomatic ones is in line with the previous findings suggesting that asymptomatic carriers have a weaker humoral immune response to COVID-19 infection (Long et al., 2020). Similarly, seropositivity rate was statistically significantly higher in those with compatible CT scan findings, indicating that the severity of the disease is positively correlated with the potency of immunity. Such a finding is similar to that were reported for SARS-CoV and MERS-CoV, for whom higher levels of IgM and IgGs have also been shown to be correlated with the severity of disease (Alshukairi et al., 2016; Lee et al., 2006; Zhang et al., 2020; Zhao et al., 2020).

Although overall seropositivity among HCWs was calculated as 2.7% in our study, fluctuations between institutions were also noted. A lower rate at Farabi (0.3%) might be presumable, as PCR confirmed HCW rate was also not that high (0.5%). On the other hand, between two institutions with comparable PCR confirmed HCW rates, observed seropositivity of HCWs from Cerrahpasa was found to be significantly higher than those from UEAH (7.2% and 1.5% respectively). As we compared the work schedules of HCWs in three hospitals, we noticed that physicians in Cerrahpasa was assigned to the pandemic clinics on daily basis. On the other days of the week the same physician served at non-pandemic clinic, too. The work schedule was made on monthly basis in UEAH and Farabi. Moreover, at Cerrahpasa as being a distinguished medical school, teaching on ward rounds with medical students continuous took place until 18^th^ of March. UEAH and Farabi do not teach undergraduate students. Cerrahpasa’s buildings were being reconstructed when the outbreak began in Turkey and this might have also impaired some infection control measures.

In 21.8% of PCR-confirmed COVID-19 patients, IgG titers were found to be below 1.4 S/C which was denoted as the cut-off value of positivity by the manufacturer. Previous studies with RT-PCR positive patients have only enrolled patients with clear clinical findings (Boise et al., 2020). It is possible that the cut off might be too stringent for asymptomatic patients. Thus, a number of asymptomatically recovered HCWs with COVID-19 in high-risk group might be also remained undetected in serodiagnosis. If the cut-off was set to 1.0 S/C the seropositivity in high risk group would have increased from 2.9% to 3.9%.

The information for SARS-CoV-2 transmission among health care workers could help for the revision of health policies and immunization strategies in hospitals for a possible resurgence of the outbreak. Further, extensive knowledge of antibody seroconversion and characterization of antibody profiles throughout SARS-CoV-2 infection could provide insights for the identification of potentially targeted neutralizing antibodies.

### Limitations

Even though a significant number of employees were tested, not all of the invited HCWs in these hospitals participated in the study. Screening larger cohorts from hospitals could serve more information to monitor the course of pandemic among HCWs.

## METHODS

### Study Design and Participants

This study was conducted in three pandemic hospitals in Istanbul and Kocaeli, including University of Health Sciences Umraniye Teaching and Research Hospital (UEAH), Istanbul University-Cerrahpasa, Cerrahpasa Medical Faculty Hospital (Cerrahpasa), Darica Farabi Teaching and Research Hospital (Farabi). HCWs were invited to participate in the study. Exposure risk of the HCWs was determined by the working areas they were assigned during the pandemic. Some HCWs were on administrative leave due to their medical conditions and they were classified as having “no risk”. HCWs working in the clinics, which were kept clear of COVID-19 or had no direct contact with any patient, were classified as having “low risk”. HCWs employed in hot zones for COVID-19 transmission including emergency unit, intensive care unit (ICU), pandemic clinics, COVID outpatient clinics, COVID testing labs, departments of Infectious Diseases and Chest Medicine, and radiology department (where CT scans and chest X-rays were performed) were classified as having “high risk”. HCWs with administrative roles for supervising hot zones with regular visits were also classified in the “high risk” group. We also recruited HCWs who were diagnosed with COVID-19 at least 14 days before enrollment for the study. This group was defined as the “PCR positive” group. Demographics data, comorbidities, drug history, date of COVID-19 diagnosis, past PCR tests, results of chest computed tomography (CT) scans were noted. HCWs not willing to give consent, HCWs with a history of COVID-19 diagnosis without a confirmatory PCR test and those diagnosed within the last 14 days were excluded. All blood samples were collected in the three hospitals in between 30^th^ May and 6^th^ of June 2020. Oro-nasopharyngeal swab samples of the three subgroups(excluding PCR positive group) were also tested to confirm the absence of active infection at the time when blood specimens were collected.

All sera samples were aliquoted after centrifugation of peripheral blood tubes at 800xg for 12 minutes and sera samples were kept in –20ºC until the study day. For detection of SARS-CoV-2 IgG, chemiluminescent microparticle immunoassay was carried out according to manufacturer’s instructions and samples were run on the related instrument (ARCHITECT, Abbott Laboratories, Abbott Park, IL, USA). Minimum 100 µL of serum was required for analysis. Qualitative results were reported by the instrument with the cut-off value of 1.40 S/C as recommended. This study was approved by the ethics committee of the Umraniye Teaching and Research Hospital (approval number: 29.05.2020/10337). Written informed consent was obtained from each enrolled participant. All study was carried out in accordance with the ethical standards of the Helsinki Declaration.

### Statistical Analysis

All statistical analyses were performed by SPSS version 22 software (Chicago, IL). Parametric variables were analyzed by Student’s *t*-test. Mann-Whitney U test was employed for nonparametric continuous variables. Categorical variables were tested by using the chi-square and Fisher’s exact test. Two-sided *p* value below 0.05 was considered to be statistically significant.

## Data Availability

Supporting information is available in supplementary files. No additional data are available.

## ACKNOWLEDGMENTS

We appreciate the generous contribution of Abbott for providing Architect SARS-CoV-2 IgG antibody kits, the chemiluminescence microparticle immunoassay method, to our institutions. We thank Yesim Karaca, Aleyna Karaca, İrem Yaren Nur and Murat Kaya for technical assistance.

## AUTHOR CONTRIBUTIONS

GA was responsible for the analysis and interpretation of the data, drafting the manuscript and literature search. AhM was responsible for data acquisition, analysis, drafting the manuscript and literature search. ZA participated to the data acquisition, literature search and study design. OT was responsible for interpreting the data, drafting the manuscript and searching the literature. NBA, AI and MG collected data and participated to the analysis. MA and RK were responsible for data acquisition. BY contributed to the design and coordination of the study. HT and AdM coordinated and operated the study. MD interpreted the collected data. FB and NS collected data, designed and coordinated the study. GDD contributed to the study design and manuscript drafting. LD was the coordinator of the project and was responsible for acquisition, interpretation and analysis of the data and drafting the manuscript. All authors critically reviewed the draft and approved the final version for publication.

## SUPPLEMENTARY DATA

**Figure S1:** Result of IgG titration means according to risk groups

**Table S1:** IgG titration values of “PCR positive” group

**Table S2:** IgG titration values of seropositive non-infected HCWs

